# Contralateral Neck Recurrence Rates After Ipsilateral Neck Adjuvant Radiation in Head and Neck Carcinomas with a Pathologically Negative Contralateral Neck

**DOI:** 10.1101/2024.05.16.24307485

**Authors:** Kendall J. Kiser, Anthony J. Apicelli, Randall J. Brenneman, Jessika A. Contreras, Hiram Gay, Michael J. Moravan, Nikhil Rammohan, Ryan S. Jackson, Patrik Pipkorn, Sidharth V. Puram, Jason T. Rich, Jose P. Zevallos, Douglas R. Adkins, Peter Oppelt, Wade L. Thorstad

**Affiliations:** Department of Radiation Oncology, Washington University School of Medicine in St. Louis, St. Louis, MO; Springfield Clinic Cancer Center, Springfield, IL; Department of Otolaryngology, Washington University School of Medicine in St. Louis, St. Louis, MO; Department of Otolaryngology, University of Pittsburgh School of Medicine, Pittsburgh, PA; Division of Oncology, Department of Medicine, Washington University School of Medicine in St. Louis, St. Louis, MO

## Abstract

**Introduction:** In 2019 we published a phase II trial of ipsilateral neck adjuvant radiation therapy in head and neck (HN) carcinoma patients with a pathologically negative (pN0) contralateral neck after bilateral neck dissection. The five-year contralateral neck recurrence rate was 3%. Here we present the recurrence rate in patients subsequently treated by this trial’s paradigm.

**Methods:** Patients were selected who would have been eligible for our phase II trial: all had oropharynx, hypopharynx, oral cavity, or unknown primary carcinomas without prior HN cancer or major HN surgeries, underwent primary resection including bilateral neck dissection with a pN0 contralateral neck, and underwent adjuvant intensity modulated radiation therapy (IMRT) that spared the contralateral neck.

**Results:** Fifty-five patients met cohort inclusion criteria. Thirty-nine cancers arose from the oropharynx, 11 from the oral cavity, three from the hypopharynx, and two from an unknown primary. With a median follow-up of 15 months there were nine recurrences (16%), four contralateral neck recurrences (7%), and one isolated contralateral neck recurrence (2%). No contralateral neck recurrences occurred in patients with p16+ oropharyngeal primaries despite that most (73%) arose from the base of the tongue. In contrast, three contralateral neck recurrences occurred in patients with advanced stage oral cavity primaries.

**Conclusion:** The contralateral neck recurrence rate in HN carcinoma patients with a pN0 contralateral neck treated by adjuvant ipsilateral neck IMRT was similar to the 3% rate observed in our phase II trial. However, this paradigm may be inadequate for patients with locally advanced oral cavity primaries.

## Introduction

In 2019, we reported the five-year outcomes of a phase II institutional trial (NCT00593840) wherein 73 patients with squamous cell carcinomas of the oral cavity (OC), oropharynx (OPX), hypopharynx, larynx, or unknown primary underwent ipsilateral-only adjuvant radiation therapy (RT) after surgical resection and bilateral neck dissection with a pathologically node-negative (pN0) contralateral neck.^1^ The trial accrued from 2007 through 2014 and predated the American Joint Commission on Cancer (AJCC) 8th edition staging system that distinguishes between p16+ and p16-OPX cancers, but by AJCC 7th edition staging 78% of patients were stage IV and 71% of patients had midline involvement. The hypothesis was that contralateral neck control would be greater than 90%, and the primary endpoint was the recurrence rate in the unirradiated pN0 neck. With a median follow-up of 53 months, just two patients (3%) experienced a contralateral neck recurrence.

Following that trial’s analysis, our institution adopted ipsilateral neck-only RT as a standard for patients who would meet trial inclusion criteria. It is important to realize that patients with lateralized tonsil cancer (primary > 1 cm from midline) were not eligible for this trial, as the institutional standard was to treat only the ipsilateral neck in these patients, including those with pN2b disease. Multiple retrospective and prospective studies have demonstrated low contralateral neck recurrence for patients with lateralized primary tonsil cancer who receive neither surgery nor elective radiation therapy.^2-7^

We now have several years of data prospectively treating patients by the phase II trial paradigm, and we undertook the present retrospective study to determine if the results from the original study could be replicated in patients treated subsequently. The present study hypothesized that neck control in the unirradiated pN0 neck would remain greater than 90%.

## Materials and Methods

### Patients

Cohort identification began by extracting all head and neck (HN) intensity modulated radiation therapy (IMRT) order sets entered from 2020-2024 (n = 855). A radiation oncologist reviewed the IMRT plans individually and categorized the neck targets and prescription dose distributions as unilateral (n = 302) or bilateral (n = 530). The radiation oncologist then reviewed electronic health records of unilateral neck RT patients to refine the patient cohort to OPX, OC, hypopharynx, larynx, or unknown primary cancers (n = 164) who underwent upfront surgical treatment with a neck dissection (n = 140) that was bilateral (n = 61) and pathologically without nodal involvement in the contralateral neck (n = 60). Five additional patients were excluded as they would not have met eligibility for the phase II trial (prior major HN surgery (n = 2), prior H&N cancer (n = 1), palliative intent RT (n = 1), and treatment refusal after 5/33 prescribed fractions (n = 1)). The final cohort was 55 patients. This study was approved by an institutional IRB (#202305057).

### Statistics

Clinical, radiographic, and laboratory variables were extracted for cohort patients. Patient variable distributions were visualized, and continuous variable distributions were assessed for approximate normality (−1 ≤ skew ≤ 1 and −2 ≤ kurtosis ≤ −2). As appropriate for each variable’s distribution, variables were evaluated with log-rank, independent T, or Mann-Whitney U tests for associations with contralateral neck recurrence outcomes. A subset of variables with statistically significant univariate associations with contralateral neck recurrence and with clinical utility/actionability were subsequently evaluated in a multivariable Cox proportional hazards model.^8^ Statistical tests were performed in Python with SciPy^9^ and Lifelines^10^ code libraries.

## Results

Fifty-five patients were included in the study cohort. Patient characteristics are summarized in Table 1. The median age was 60 years (interquartile range (IQR): 55 – 68 years). Most patients were male (80%) and were current or former smokers (52%) with a median 27.5 pack-year smoking history (IQR: 10 – 37.5). Primary sites included the OPX (71%), OC (20%), hypopharynx (5%), or unknown primary (4%). All patients but one (98%) had squamous cell carcinoma histology. Immunohistochemical staining for p16 was positive in 71% of patients (92% of OPX), negative in 18% of patients (8% of OPX), and not tested in 11% of patients (all OPX were tested). Surgical margins were negative in 65% of cases, close (≤ 2 mm) in 24%, and positive in 11%. LVSI and PNI were present in 36% and 22% of cases, respectively. All patients but three (95%) underwent PET-CT staging. Among 36 p16+ OPX patients, 12 (33%) had ≥ 2 pathologically involved nodes and 12 (33%) had ENE. Among all patients, pathologic summary stages were as follows: stage I: 53%; stage II: 15%; stage III: 2%; stage IV: 30% (AJCC 8^th^). Further clinical characteristics are available in Supplementary Table 1.

**Table 1:**
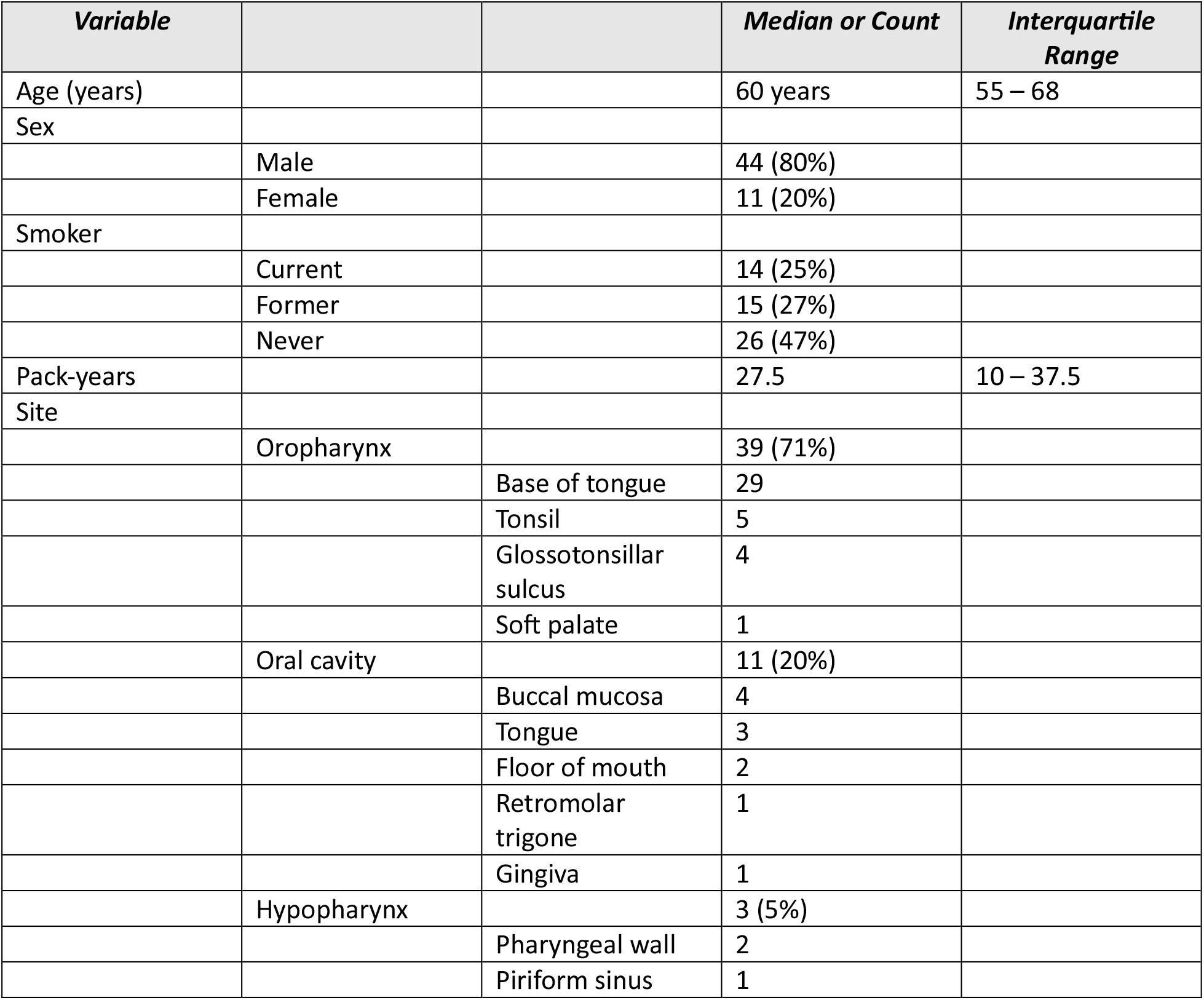
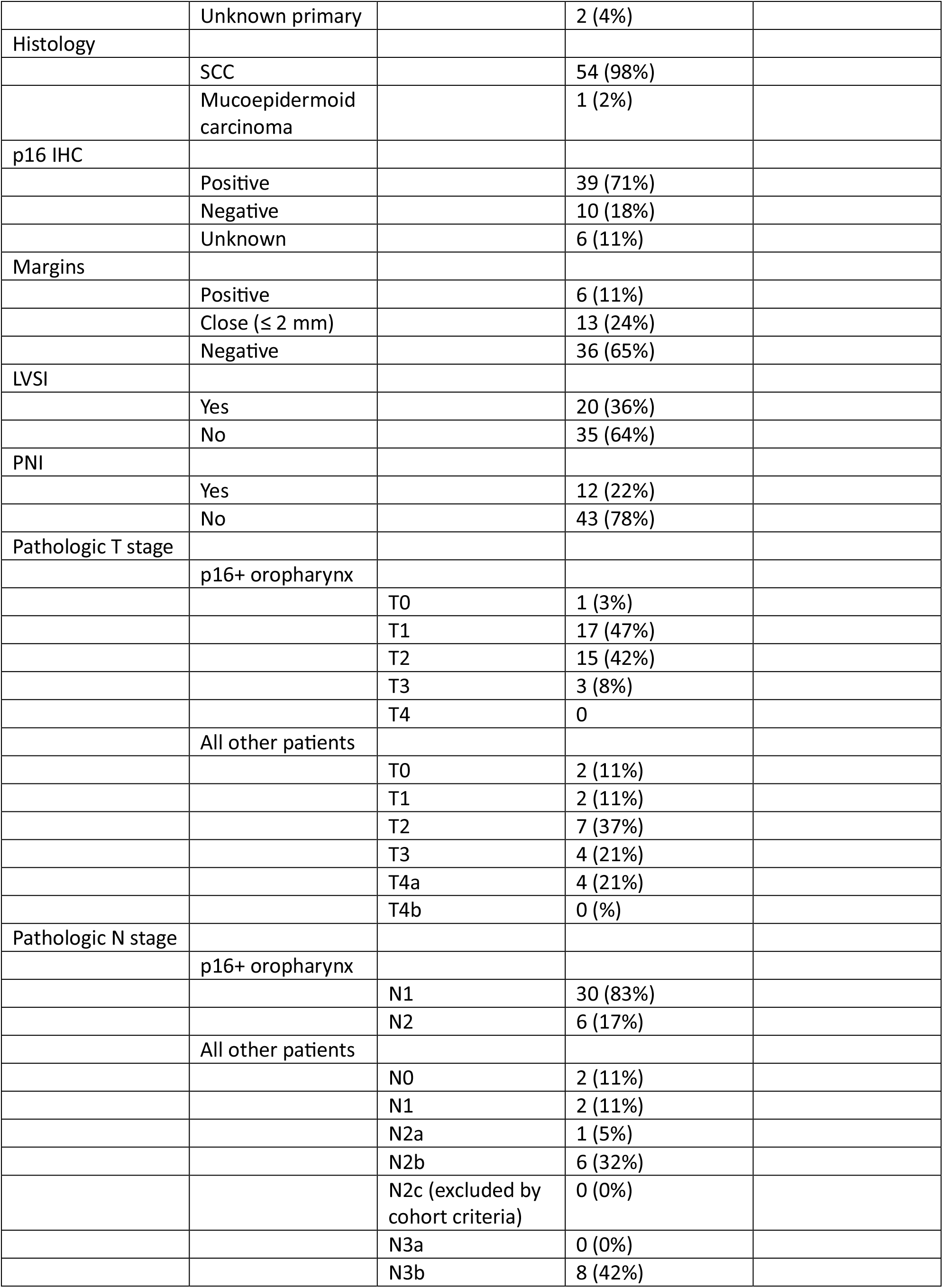
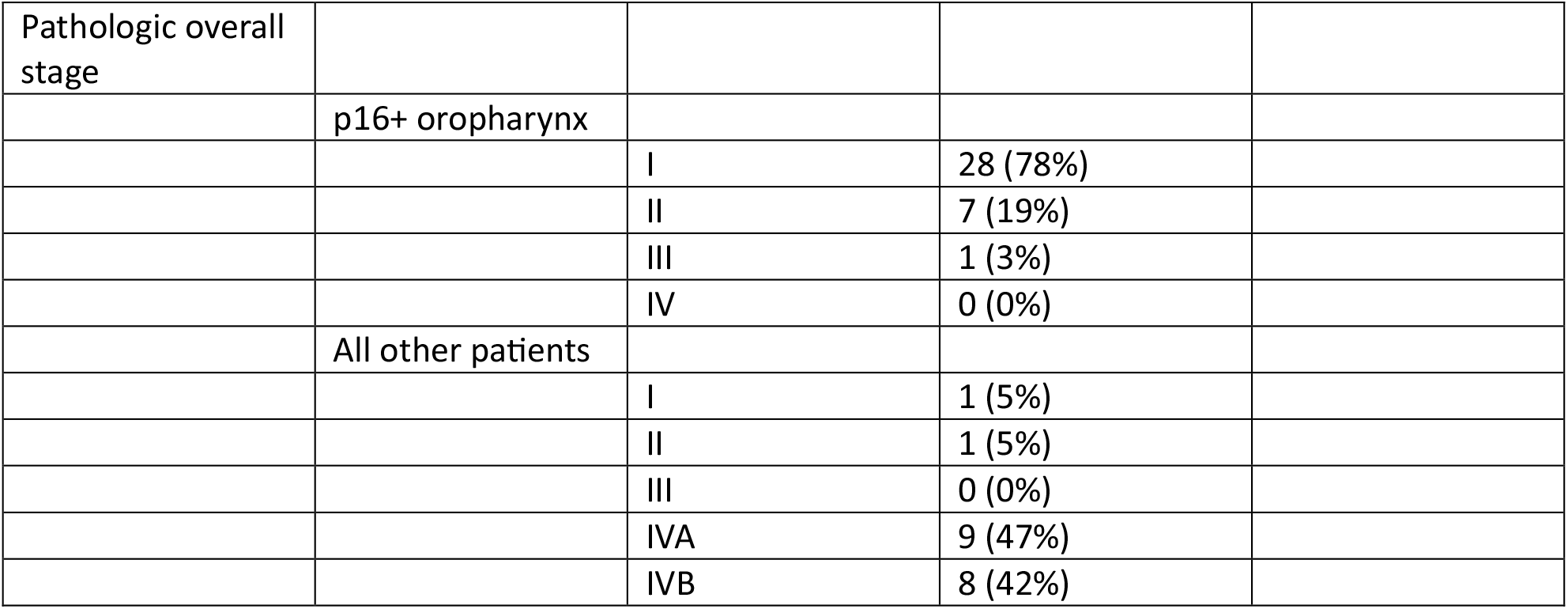
Patient characteristics.

**Table 1:**
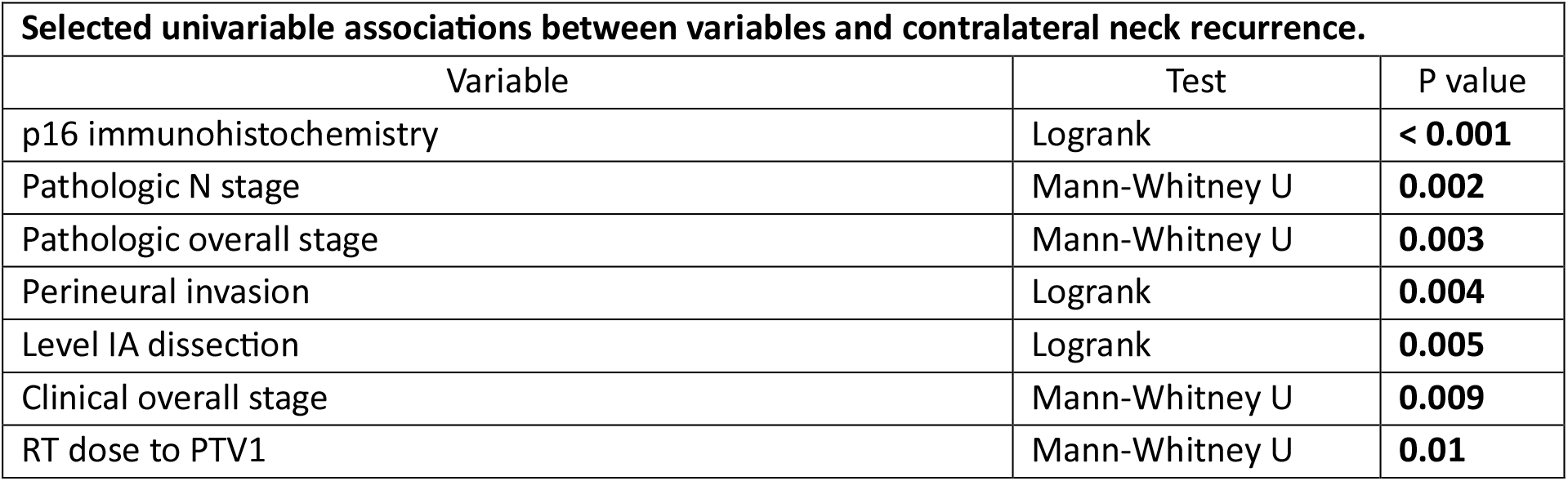
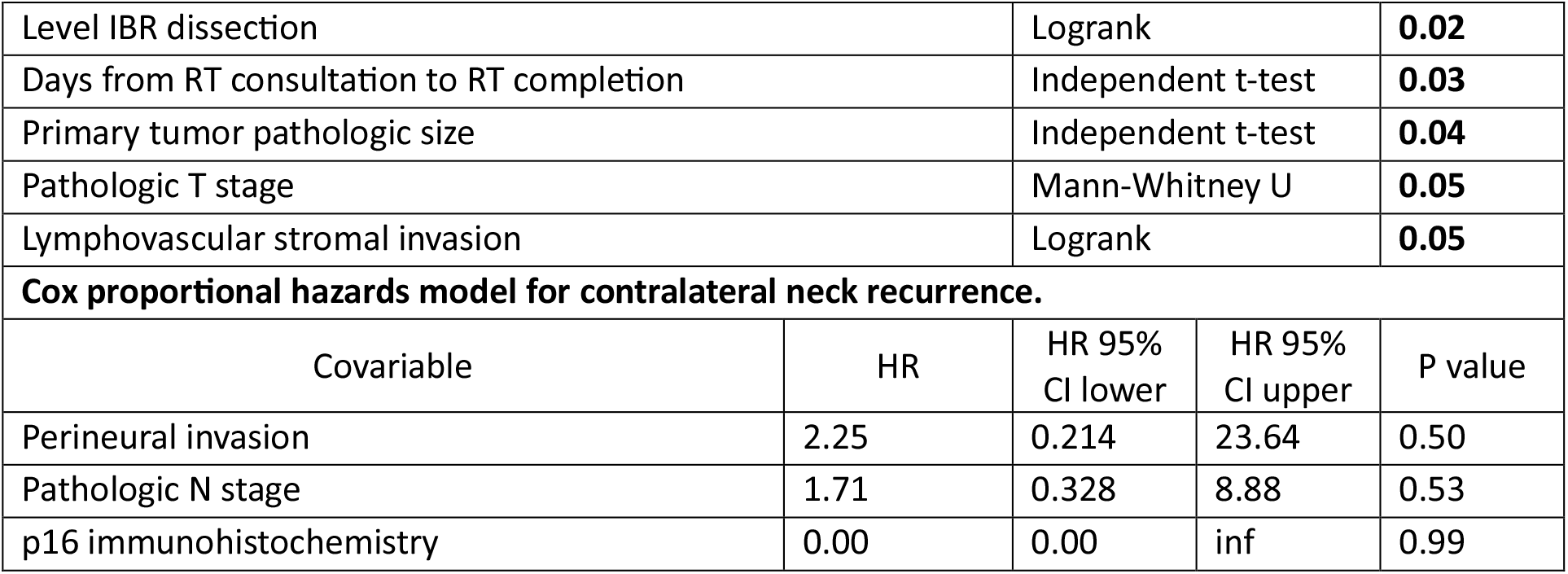
Univariable and multivariable associations between selected variables.

| Selected univariable associations between variables and contralateral neck recurrence. |  |  |
| --- | --- | --- |
| Variable | Test | P value |
| p16 immunohistochemistry | Logrank | **< 0.001** |
| Pathologic N stage | Mann-Whitney U | **0.002** |
| Pathologic overall stage | Mann-Whitney U | **0.003** |
| Perineural invasion | Logrank | **0.004** |
| Level IA dissection | Logrank | **0.005** |
| Clinical overall stage | Mann-Whitney U | **0.009** |
| RT dose to PTV1 | Mann-Whitney U | **0.01** |

|  |  |  |  |  |
| --- | --- | --- | --- | --- |
| Level IBR dissection | Logrank | **0.02** |  |  |
| Days from RT consultation to RT completion | Independent t-test | **0.03** |  |  |
| Primary tumor pathologic size | Independent t-test | **0.04** |  |  |
| Pathologic T stage | Mann-Whitney U | **0.05** |  |  |
| Lymphovascular stromal invasion | Logrank | **0.05** |  |  |
| **Cox proportional hazards model for contralateral neck recurrence.** |  |  |  |  |
| Covariable | HR | HR 95%<br>CI lower | HR 95%<br>CI upper | P value |
| Perineural invasion | 2.25 | 0.214 | 23.64 | 0.50 |
| Pathologic N stage | 1.71 | 0.328 | 8.88 | 0.53 |
| p16 immunohistochemistry | 0.00 | 0.00 | inf | 0.99 |

For p16+ OPX cases, adjuvant RT prescriptions were most commonly 60 Gy to the primary and involved nodal planning target volumes (PTVs) and 52 Gy to the elective ipsilateral neck. In contrast, for p16-OPX and other primaries the prescriptions were usually 66 Gy to the primary and involved nodal PTVs and 54 Gy to the elective ipsilateral neck. A minority of cases (13%) were planned and treated with three dose levels. Twenty-five percent of courses were delivered with concurrent chemotherapy (cisplatin in all but one case), and percutaneous gastrostomy tubes were placed in 24% of patients during treatment. Time intervals between clinical events are visualized in supplementary figure 1.

With a median follow-up of 15 months (range: 2 – 38), the number of events and event rates of any recurrence, contralateral neck recurrence, or isolated contralateral neck recurrence were respectively nine events (16%), four events (7%), and one event (2%). There were no contralateral neck recurrences among patients with p16+ OPX primaries (n = 36) or those with less than stage IVA disease (Figure 1A). The single isolated contralateral neck recurrence was in a patient with a p16-OPX tumor. The three remaining contralateral neck recurrences were OC tumors (Figure 1B): one had a synchronous primary recurrence, one had synchronous primary, bilateral neck, and distant disease, and one developed widespread distant disease without local or regional disease but ultimately failed in the contralateral neck 14 months later. The median time to contralateral neck recurrence was 7.6 months (IQR: 6.2 – 9.7). There were eight deaths (15%) occurring at a median of 17 months from RT consultation (IQR: 11 – 22) (Figure 1C).

**Figure 1:**
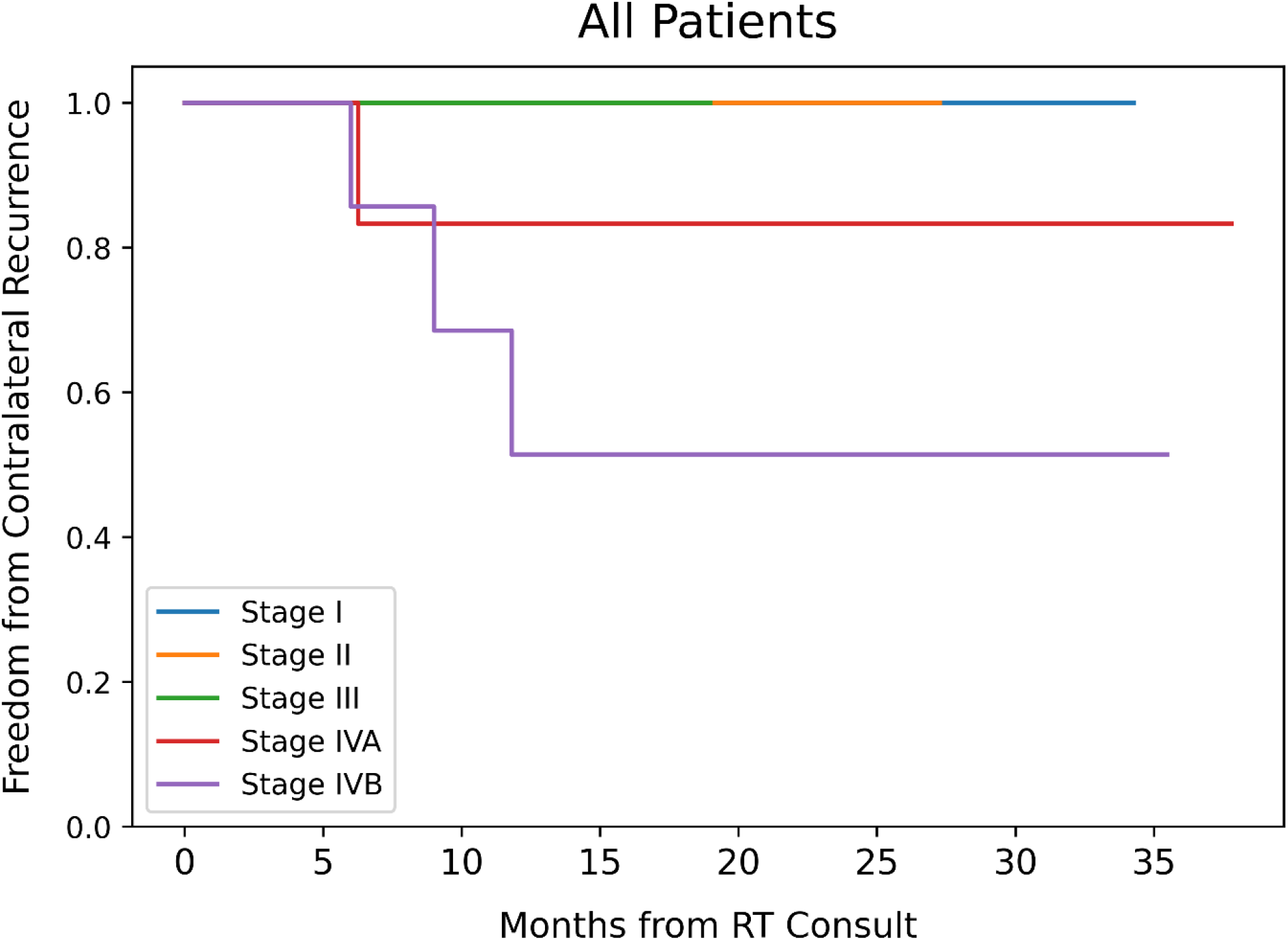
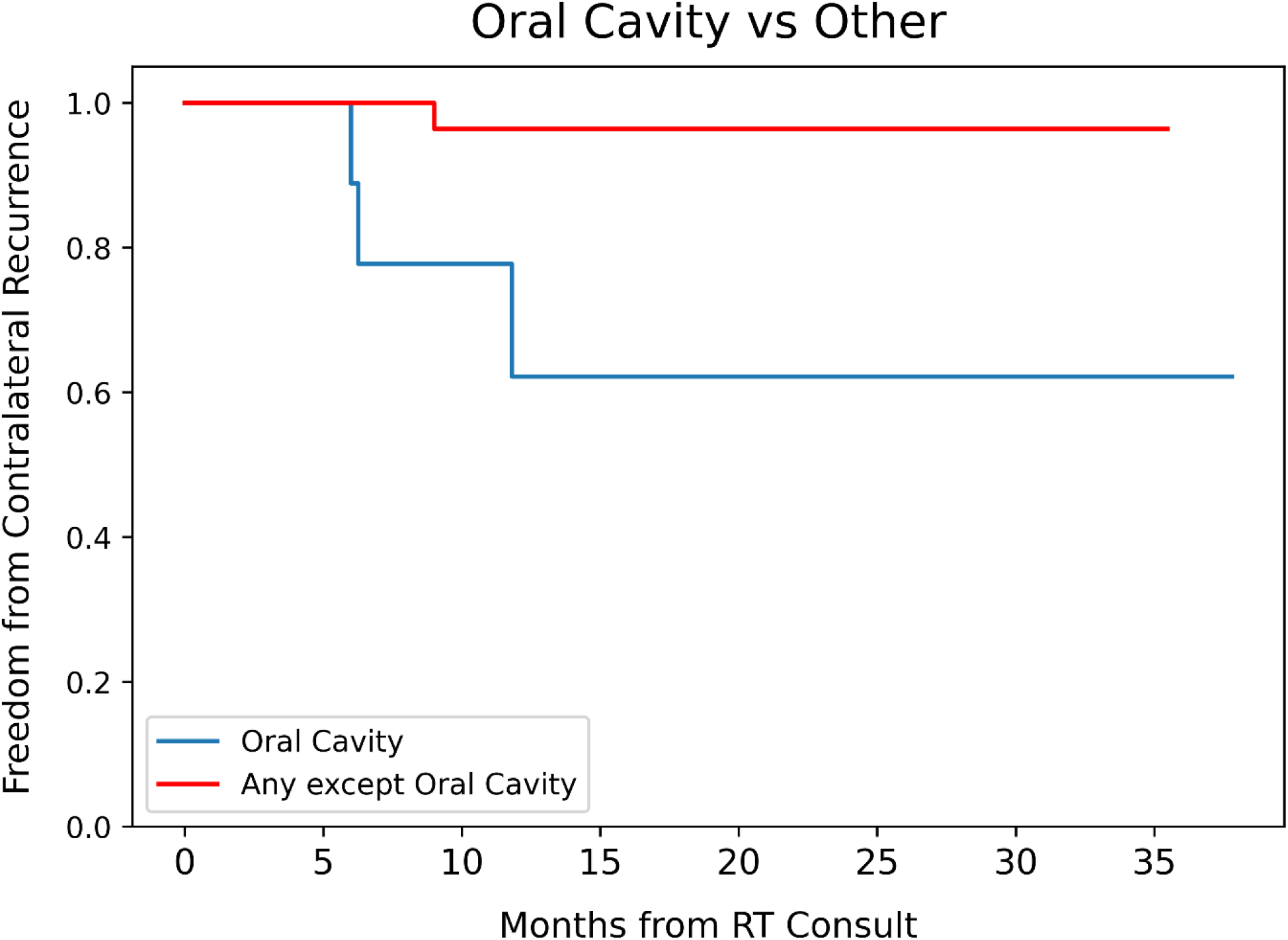
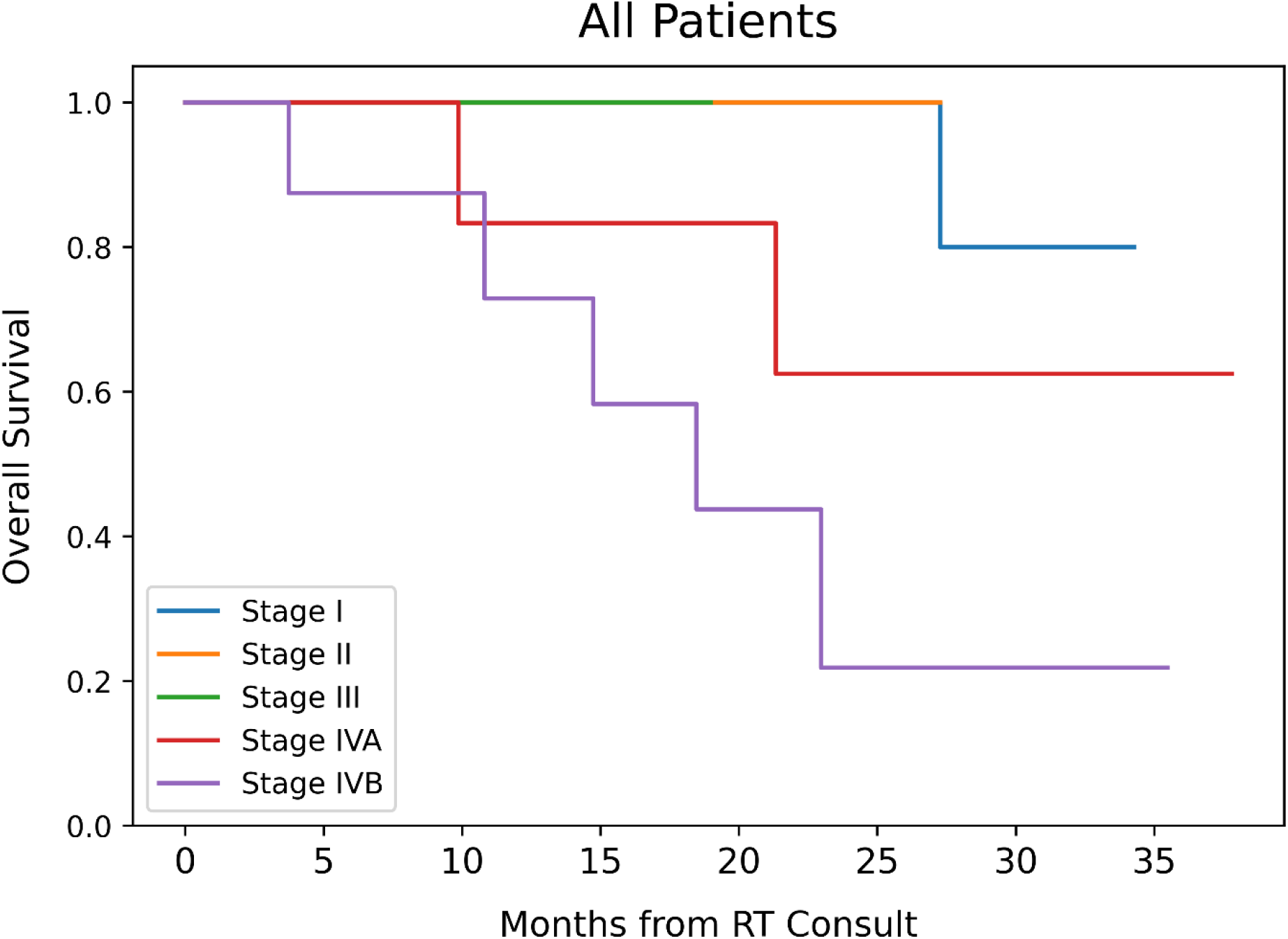
Kaplan-Meier curves visualize significantly higher contralateral recurrence rates in stage IVA or IVB disease than in earlier stages (A; logrank IVA vs. I: p = 0.06; IVB vs. I: p < 0.005). Contralateral neck recurrence rates were higher in OC primaries compared to OPX, hypopharynx, or unknown primaries (B; logrank p < 0.005). (C) visualizes patient overall survival was stratified by pathologic summary stage.

Clinical variables were tested for associations with contralateral neck recurrence. Selected univariable and multivariable associations with contralateral neck recurrence are listed in Table 2. Pathologic T, N, and summary stages, PNI, LVSI, primary tumor size, p16 status, and time from RT consultation to RT completion were among the significant univariable correlates with contralateral neck recurrences. However, no variable remained significantly associated with contralateral neck recurrence in a multivariable Cox model incorporating p16, pathologic N stage, and PNI covariables.

## Discussion

This was a retrospective review of 55 HN cancer patients with OPX, OC, hypopharynx, or unknown primaries who would have been candidates for a prior institutional phase II trial omitting contralateral neck RT^1^ and per that paradigm received adjuvant ipsilateral neck RT after an upfront surgery and bilateral neck dissections that resulted in a pN0 contralateral neck. The contralateral neck recurrence rate was similar (7%) to the 5-year contralateral neck recurrence rate in that trial (3%) despite shorter median follow-up time. Contralateral neck recurrences occurred in three patients with stage IVA or IVB OC primary cancers and one patient with a p16-OPX primary. For the three OC contralateral recurrences, one was synchronous with a primary recurrence, one was synchronous with widespread metastatic disease, and one occurred 14 months after developing widespread metastatic disease. It is doubtful that these contralateral recurrences impacted the patients’ subsequent clinical courses. The patient with the p16-OPX cancer had an isolated contralateral recurrence, and this impacted the subsequent clinical course.

To frame these results, two recent meta-analyses of ipsilateral neck RT for tonsillar^11^ and OC^12^ cancers published by Razavian et al. are instructive. In the first^11^ – a meta-analysis of ipsilateral neck RT delivered to 1487 tonsillar cancer patients drawn from 17 studies – the pooled rate of contralateral neck recurrence was 1.9%. Increasing T stages were associated with higher contralateral recurrence risk (up to 16% for T4 tumors) but increasing N stages (N2b – N3 vs. N0 – N2a) were not (3% vs. 1.7%, p = 0.07). In the subset of studies that compared ipsilateral with bilateral neck RT, bilateral RT was associated with a slightly lower risk of contralateral recurrence (2.8% vs. 0.9%, p = 0.04). Taku et al.^13^ published after this meta-analysis’s data collection period ended, but their data further corroborates a low contralateral neck recurrence rate (2.2%) from a retrospective cohort of 403 patients with T1-2N0-2b well-lateralized tonsillar primaries who were treated with ipsilateral neck 3D-CRT, IMRT, or protons between 2000 and 2018 and followed for a median of 5.8 years.

In the second Razavian manuscript^12^ – a meta-analysis of ipsilateral neck RT delivered to 1825 OC cancer patients drawn from 15 studies – the pooled rate of contralateral neck recurrence was 5.7% and not significantly different between OC subsites. Unlike OPX, increasing T and N stages were associated with a significantly higher risk of contralateral neck recurrence (T3-T4: 9%; N2-N3: 17.4%). Of the 41 patients who progressed in the contralateral neck and for whom pT and pN staging data were available, 80% had both pT4 and pN2-N3 disease.

Our findings may extend the criteria of which p16+ OPX patients can be considered for ipsilateral neck RT. First, prior studies excluded OPX patients with base of tongue or midline involvement. Indeed, the 2020 American Radium Society guideline for ipsilateral neck RT in tonsillar cancers^14^ (an update of the 2011 American College of Radiology appropriate use criteria^15^) recommends “use of ipsilateral neck irradiation as usually appropriate after resection of a well-lateralized tonsil primary tumor with a single ipsilateral pathologically positive node.” However, our study found no recurrences in the pN0 contralateral neck in a p16+ OPX population despite most cases arising from the base of tongue (73%). Importantly, prior studies have not always reported or required a bilateral neck dissection.

Second, to our knowledge,^11,12^ this is just the second study^16^ of ipsilateral neck RT to reference AJCC 8^th^ edition staging and the first to use it for an OPX cohort. The American Radium Society guideline comments, “The literature review performed for this guideline did not return any titles that used the AJCC 8^th^ edition staging system.” Perhaps for inability to separately evaluate HPV+ oropharynx nodal staging, the guideline “strongly recommends the use of bilateral neck irradiation as usually appropriate after neck dissection in cases of multiple pathologically positive ipsilateral lymph nodes and in the presence of macroscopic extranodal extension.” However, 29% of our p16+ OPX population had ≥ 2 pathologically positive nodes and 32% had ENE. Using the updated 8^th^ edition staging, no p16+ OPX patient had worse than stage III disease, whereas formerly many of these patients would have been upstaged to stage IV disease by nodal involvement.

For patients with OC primaries “at substantial risk of contralateral nodal involvement – for example, tumor of the oral tongue and/or floor of the mouth that is T3 or T4,” the American Society of Clinical Oncology Clinical Practice Guidelines for the Management of the Neck^17^ recommend bilateral adjuvant neck RT if the patient received only an ipsilateral neck dissection. These guidelines do not advocate for or against bilateral neck RT following a bilateral neck dissection. It is of interest that three of the four contralateral neck recurrences in the present study and two (out of two) in the phase II study were from OC primary tumors, raising some concern about omitting contralateral radiation therapy for locally advanced OC primary tumors. However, none of these five OC neck recurrences were isolated; they all were part of synchronous and/or distant recurrences. The phase II PRESERVE trial is currently accruing patients with T1-4 N0-2 OC cancers with at least one pN0 hemi-neck to a randomization between standard RT volumes or omission of RT in the pN0 hemi-neck. This trial’s primary endpoint – regional failure in the pN0 hemi-neck – will provide further guidance for this cohort in coming years.^18^

## Conclusion

The results of this study corroborate our initial phase II trial showing that omission of RT to the contralateral pN0 neck results in a control rate of greater than 90%. There were no contralateral neck recurrences in p16+ OPX primaries or those with less than stage IVA disease. However, this approach may not be appropriate for locally advanced OC primary tumors as three of the four contralateral neck recurrences occurred in patients with stage IVA or IVB OC disease.

## Data Availability

All data produced in the present study are available upon reasonable request to the authors

