## Supplementary Table 1 for "Contralateral Neck Recurrence Rates After Ipsilateral Neck Adjuvant Radiation in Head and Neck Carcinomas with a Pathologically Negative Contralateral Neck"

| Staging FDG-PET |  |  |  |  |
| --- | --- | --- | --- | --- |
|  | Primary SUV max |  | 11.7 | 8.9 – 16.6 |
|  | Node SUV max |  | 11.1 | 7.5 – 14.5 |
| Pathologic primary size |  |  | 2.2 cm | 1.6 – 3.4 cm |
| Depth of invasion |  |  | 1.2 cm | 0.9 – 1.8 cm |
| Grade |  |  |  |  |
|  | 2 |  | 10 (18%) |  |
|  | 3 |  | 5 (9%) |  |
|  | none |  | 40 (73%) |  |
| Dissected nodal levels |  |  |  |  |
|  | IA |  | 11 (20%) |  |
|  | IBR |  | 13 (24%) |  |
|  | IBL |  | 12 (22%) |  |
|  | IIAR |  | 54 (98%) |  |
|  | IIAL |  | 54 (98%) |  |
|  | IIBR |  | 46 (84%) |  |
|  | IIBL |  | 47 (85%) |  |
|  | IIIR |  | 54 (98%) |  |
|  | IIIL |  | 52 (95%) |  |
|  | IVR |  | 52 (95%) |  |
|  | IVL |  | 48 (87%) |  |
|  | VR |  | 1 (2%) |  |
|  | VL |  | 1 (2%) |  |
| Resected nodes |  |  |  |  |
|  | Right |  | 31 nodes | 21 – 38 nodes |
|  | Left |  | 25 nodes | 17 – 34 nodes |
| Positive nodes |  |  |  |  |
|  | Ipsilateral side |  | 2 nodes | 1 – 3 nodes |
| Involved level 4 or 5 |  |  |  |  |
|  | Yes |  | 11 (20%) |  |
|  | No |  | 44 (80%) |  |
| Largest node size |  |  | 3.5 cm | 1.6 – 4.5 cm |
| ENE |  |  |  |  |
|  | Yes |  | 20 (36%) |  |
|  | No |  | 35 (64%) |  |
| Tumor genomic sequencing |  |  |  |  |
|  | Yes |  | 8 (15%) |  |
|  | No |  | 47 (85%) |  |
| Radiation dose level 1 |  |  |  |  |
|  | Patients |  | 55 (100%) |  |
|  | Dose |  | 60 Gy | 60 – 66 Gy |
| Radiation dose level 2 |  |  |  |  |
|  | Patients |  | 46 (84%) |  |
|  | Dose |  | 52 Gy | 52 – 54 Gy |
| Radiation dose level 3 |  |  |  |  |
|  | Patients |  | 7 (13%) |  |
|  | Dose |  | 52 Gy | 52 – 54 Gy |
| Radiation course duration |  |  | 43 days | 41 – 46.5 days |
| Concurrent Chemotherapy |  |  |  |  |
|  | Cisplatin |  | 13 (24%) |  |
|  |  | dosed 100 mg/m^2^ q 3 weeks | 8 |  |
|  |  | dosed 40 mg/m^2^ weekly | 5 |  |
|  | Cetuximab/Docetaxel |  | 1 (2%) |  |
|  | None |  | 41 (75%) |  |
| Percutaneous gastric tube |  |  |  |  |
|  | Yes |  | 13 (24%) |  |
|  | No |  | 42 (76%) |  |
| Clinical T stage |  |  |  |  |
|  | p16+ oropharynx |  |  |  |
|  |  | T0 | 9 (25%) |  |
|  |  | T1 | 16 (44%) |  |
|  |  | T2 | 10 (28%) |  |
|  |  | T3 | 1 (3%) |  |
|  |  | T4 | 0 (0%) |  |
|  | All other patients |  |  |  |
|  |  | T0 | 2 (11%) |  |
|  |  | T1 | 5 (26%) |  |
|  |  | T2 | 4 (21%) |  |
|  |  | T3 | 4 (21%) |  |
|  |  | T4a | 4 (21%) |  |
|  |  | T4b | 0 (0%) |  |
| Clinical N stage |  |  |  |  |
|  | p16+ oropharynx |  |  |  |
|  |  | N1 | 34 (94%) |  |
|  |  | N2 | 2 (6%) |  |
|  |  | N3 | 0 (0%) |  |
|  | All other patients |  |  |  |
|  |  | N0 | 6 (32%) |  |
|  |  | N1 | 3 (16%) |  |
|  |  | N2a | 0 (0%) |  |
|  |  | N2b | 7 (37%) |  |
|  |  | N2c | 1 (5%) |  |
|  |  | N3a | 0 (0%) |  |
|  |  | N3b | 2 (11%) |  |
| Clinical overall stage |  |  |  |  |
|  | p16+ oropharynx |  |  |  |
|  |  | I | 33 (92%) |  |
|  |  | II | 3 (8%) |  |
|  |  | III | 0 (0%) |  |
|  |  | IV | 0 (0%) |  |
|  | All other patients |  |  |  |
|  |  | I | 3 (16%) |  |
|  |  | II | 2 (11%) |  |
|  |  | III | 3 (16%) |  |
|  |  | IVA | 10 (53%) |  |
|  |  | IVB | 1 (5%) |  |
