## Supplementary Figure 1 for "Contralateral Neck Recurrence Rates After Ipsilateral Neck Adjuvant Radiation in Head and Neck Carcinomas with a Pathologically Negative Contralateral Neck"

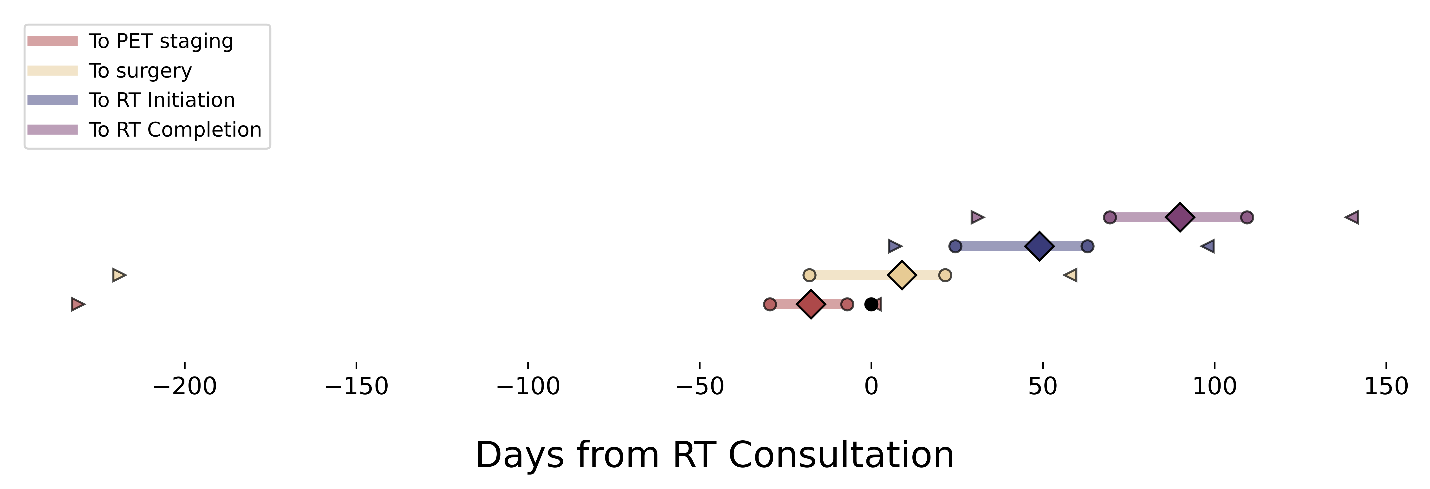

*Supplementary Figure 1:* Time intervals for PET/CT, surgery, RT first fraction and RT final fraction relative to the RT consultation date. Diamonds plot median number of days, circles plot interquartile ranges, and triangles plot the most extreme (fewest or most) number of days from RT consultation to each respective event. The median time from RT consultation to the staging PET scan was -18 days (IQR: -29 – -8). The median time from RT consultation to surgery was nine days (IQR: -18 – 22). The median time from RT consultation to the first fraction was 49 days (IQR: 25 – 63) and to the final fraction was 90 days (IQR: 70 – 110). RT courses lasted a median of 43 days (IQR: 41 – 47).
